# No Field for Lactation: Inferring Lactation Status From US Commercial Claims to Describe Postpartum Medication Dispensing

**DOI:** 10.64898/2026.09.23.26363728

**Authors:** Kaytlin Krutsch, Duke Appiah

**Affiliations:** InfantRisk Center of Excellence, Texas Tech University Health Sciences Center, Amarillo, Texas, USA; Department of Obstetrics and Gynecology, School of Medicine, Texas Tech University Health Sciences Center, Amarillo, Texas, USA; Department of Public Health, Julia Jones Matthews School of Population and Public Health, Texas Tech University Health Sciences Center, Lubbock, Texas, USA

**Keywords:** lactation, breastfeeding, pharmacoepidemiology, administrative claims data, drug utilization, postpartum period, phenotype

## Abstract

**Purpose:** Lactation status is not recorded as a discrete data element in U.S. administrative claims databases, limiting research on lactation in health databases. We developed a claims-based phenotype to identify periods of probable lactation and used it to describe medications dispensed to lactating women enrolled in a national health plan.

**Methods:** This retrospective cohort study used data from the 2012-2014 Merative MarketScan Commercial Claims Database. Deliveries were identified by diagnosis-related group classification system. Lactation windows ran from the delivery date to the date of the last surrogate lactation indicator, such as breast pump and supply codes, the encounter code for supervision of a lactating mother, and mastitis. New prescriptions filled within a window were characterized by therapeutic group, prescriber type, lactation risk category, and availability of published lactation data.

**Results:** Of 566,284 women with a single delivery, 84,679 were determined to be lactating using the developed algorithm based on maternal phenotype. The median length of assignable lactation window was 17 days (IQR 5-47). Windows most often closed on a breast pump claim (53.2%) or a postpartum visit code (28.2%). Among 158,471 new prescriptions filled during lactation, central nervous system agents (54.5%), anti-infectives (17.5%), and hormonal products accounted for > 80%. Combined oral contraceptives represented 8.2% of hormonal prescriptions during lactation against 29.0% among women of unknown status. Obstetricians wrote approximately one third of prescriptions filled during lactation windows.

**Conclusions:** Lactation can be identified in administrative claims using codes already present in the data, making national description of dispensing during lactation possible.

## 1. Introduction

Postpartum women are the most poorly served population in clinical pharmacology. A 2026 landscape analysis of the maternal and pediatric pharmacology literature found that 51.4% of drugs had no evidence, or only weak evidence, across pharmacokinetic, pharmacoepidemiologic, and clinical trial study types in postpartum women. This gap was the largest of any subpopulation examined, and substantially larger than the gap for pregnant women (32.5%) or for infants under one year of age (25.8%).^1^ Roughly a third of current drug labels carry information for nursing mothers.^2^ Evidence is thinnest of all in the weeks immediately following delivery, when maternal enzyme and transporter activity are still returning toward the non-pregnant state.^1^

Which drugs that evidence should be generated for is itself unknown, because what medications lactating women are prescribed or fill has not been described at national scale. The 2026 MPRINT prioritization report, assembled from NICHD-convened expert working groups, identified the absence of lactation status in administrative data and the lack of national utilization data as recurring barriers across therapeutic areas.^3^ Existing accounts rest on case reports, small prospective series, and single-service consultation cohorts.^4-7^ This constrains clinicians, who must treat the presenting conditions of lactating patients regardless of the evidence available, and constrains researchers, who cannot prioritize pharmacokinetic study without knowing what is in use.

Lactation status is not recorded as a discrete element in administrative claims. Where lactation has been studied in real-world data, investigators have depended on systems that record it directly: infant feeding responses captured in standardized flowsheets at well-child visits in a single US health system,^8^ or, in Denmark, a national child health register to which reporting of exclusive breastfeeding duration has been mandatory since 2012.^9^ Others have used data internal to a single health system,^10^ purpose-built research cohorts,^11^ or prospective registries.^12^ Each approach requires that information on lactation is reported. In the US, the only database that follows a national population across payers and care settings with information before and after delivery are often administrative claims databases. However, claims databases inherently do not collect data on self-reported or clinician-documented lactation nor are there any published methods that identify lactation in claims databases.

We developed an algorithm consisting of surrogate indicators available in administrative claims to identify time windows during which a commercially insured enrollee was probably lactating and characterize prescriptions filled during those windows.

## 2. Methods

This was a retrospective cohort study using deidentified patient-level claims from the 2012-2014 Merative MarketScan Commercial Claims Database, generated by more than 120 U.S. health plans covering employees of large employers. Reporting follows the RECORD-PE extension of the STROBE statement.^13^ Linked data were available for the primary covered individual and dependents, including eligibility, inpatient and outpatient claims and encounters, and outpatient prescription fills. Because dependents are linked to the primary beneficiary, the database permits a lactating woman to be linked to her infant; that linkage was not used in this analysis, which was scoped to maternal dispensing and does not examine infant exposure or outcomes. Data were obtained through the Merative MarketScan Dissertation Support Program.

Deliveries were identified using published MarketScan pregnancy phenotypes combining ICD-9-CM diagnosis and procedure codes, CPT codes, and diagnosis-related groups.^14^ In this database, DRG codes alone were sufficient to identify deliveries and were used exclusively. Six delivery MS-DRGs were applied: 765 and 766 (cesarean section with and without CC/MCC), 767 and 768 (vaginal delivery with sterilization and/or D&C, and with an operating room procedure), and 774 and 775 (vaginal delivery with and without complicating diagnoses). Women with more than one delivery date during the study period were excluded to prevent mismatching of delivery and lactation dates.

Lactation was identified using surrogate indicators recorded in billing data. ICD-9-CM diagnosis codes, CPT codes, and HCPCS codes were assembled to represent breast pump equipment and replacement supplies, the encounter code for supervision of a lactating mother, mastitis and breast abscess, nipple infection and cracked nipple, engorgement, galactorrhea, and lactation class attendance. Each indicates that a woman was lactating at the time the code was billed. The complete phenotype, with corresponding ICD-10-CM codes alongside ICD-9-CM, is provided in Supplementary Table S2.

The date of each lactation indicator was followed backward to the delivery date, and a lactation window was assigned from the date of delivery to the date of the last indicator observed. Time after the end of the window was treated as undefined lactation status until the next delivery episode. Window length therefore represents a minimum observed duration of lactation rather than an estimate of breastfeeding duration.

Women with no lactation indicator had no censoring date equivalent to the last lactation indicator. The distribution of lactation window lengths observed in the lactation group, specific to delivery year, was therefore superimposed on this group. Women were selected at random without replacement and assigned a follow-up window after delivery matching that distribution. Women whose only lactation indicator belonged to one of three code families dropped during analysis (galactorrhea, delayed lactation, and candidiasis of the breast or nipple) were excluded rather than reclassified, as the relationship was unclear between code and lactation status or function.

The outpatient prescription file was searched for fills occurring after delivery. A prescription was classified as new when the claim represented an original fill rather than a refill, as indicated by the refill number recorded in the file. Prescriptions filled within a lactation window were classified as filled during lactation.

Medications were matched to lactation risk categories from Hale’s Medications and Mothers’ Milk,^15^ using the 2012 edition to correspond with the study period; the most conservative category was applied to combination products. Availability of published lactation data for each drug was taken from the 2014 edition,^16^ the first to report data availability within each monograph. Because that edition postdates part of the study period, a drug recorded as having limited data in 2014 may be misclassified for earlier prescriptions.

Prescriber type and place of service are not carried in the MarketScan prescription file. These variables were added by cross-referencing the date a prescription was written against outpatient encounters on the same date and attaching the associated provider type and place of service. Linkage rates are reported with the results.

The lactation phenotype has not been validated against a chart-confirmed reference standard. No external comparator for lactation status currently exists in U.S. claims data. Validation evidence is available for one component, though it addresses a different quantity: among patients with electronic health record documentation of lactation, ICD-10-CM codes N61.0 or O91.2 had a positive predictive value of 76% (95% CI, 67.3-82.9) for probable lactational mastitis, rising to 80% (95% CI, 69.6-87.4) when an antibiotic dispensing within three days was required.^17^ That study estimates the probability that a coded case represents true mastitis among known lactating patients, not the probability that a coded case represents lactation.

Categorical variables are reported as counts and percentages. Continuous variables are reported as mean and standard deviation when normally distributed and median with interquartile range otherwise; normality was assessed by Kolmogorov–Smirnov test. Differences by lactation status were tested by chi-square for categorical variables, independent-samples t test and ANOVA for normally distributed continuous variables, and Mann– Whitney U and Kruskal–Wallis tests for non-normally distributed continuous variables. Significance was defined as an a priori α of 0.05. Analyses were conducted in SAS 9.4 (SAS Institute, Cary, NC).

## 3. Results

Of approximately 29 million enrollees, 566,284 women had a single delivery during the study period (Figure 1). A lactation indicator was present for 160,367 of them. Of these, 84,679 had an assignable lactation window; the remaining 75,688 had an indicator that could not be anchored to a delivery and were excluded. Of the 405,917 women with no lactation indicator, 1,985 were excluded because their only indicator belonged to a code family dropped during analysis, leaving 403,932 classified as unknown lactation status. Windows had a median length of 17 days (IQR 5-47) and most often closed on a claim for a breast pump or supplies (53.2%), followed by the postpartum visit code (28.2%), mastitis or a related nipple condition (11.6%), engorgement (6.3%), and lactation class attendance (0.8%) (Table 1). Fill timing tracked the indicator that closed the window, with pump-associated fills concentrated in the first two weeks and postpartum visit fills peaking near six weeks (Figure 2).

**Table 1.**
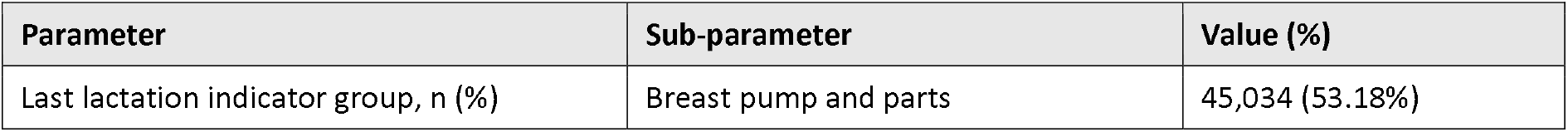
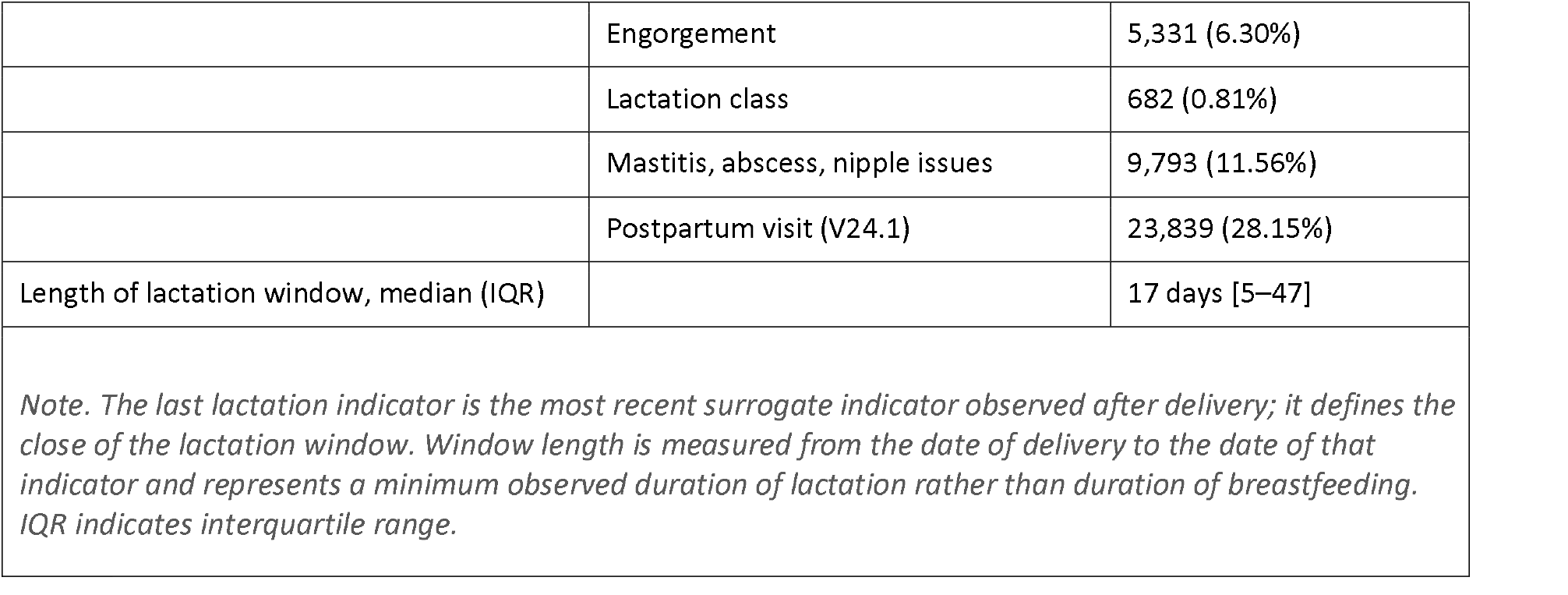
Characteristics of lactation windows among postpartum enrollees with an assignable window (N = 84,679)

| Parameter | Sub-parameter | Value (%) |
| --- | --- | --- |
| Last lactation indicator group, n (%) | Breast pump and parts | 45,034 (53.18%) |
|  | Engorgement | 5,331 (6.30%) |
|  | Lactation class | 682 (0.81%) |
|  | Mastitis, abscess, nipple issues | 9,793 (11.56%) |
|  | Postpartum visit (V24.1) | 23,839 (28.15%) |
| Length of lactation window, median (IQR) |  | 17 days [5–47] |

*Note. The last lactation indicator is the most recent surrogate indicator observed after delivery; it defines the close of the lactation window. Window length is measured from the date of delivery to the date of that indicator and represents a minimum observed duration of lactation rather than duration of breastfeeding. IQR indicates interquartile range.*

**Figure 1.**
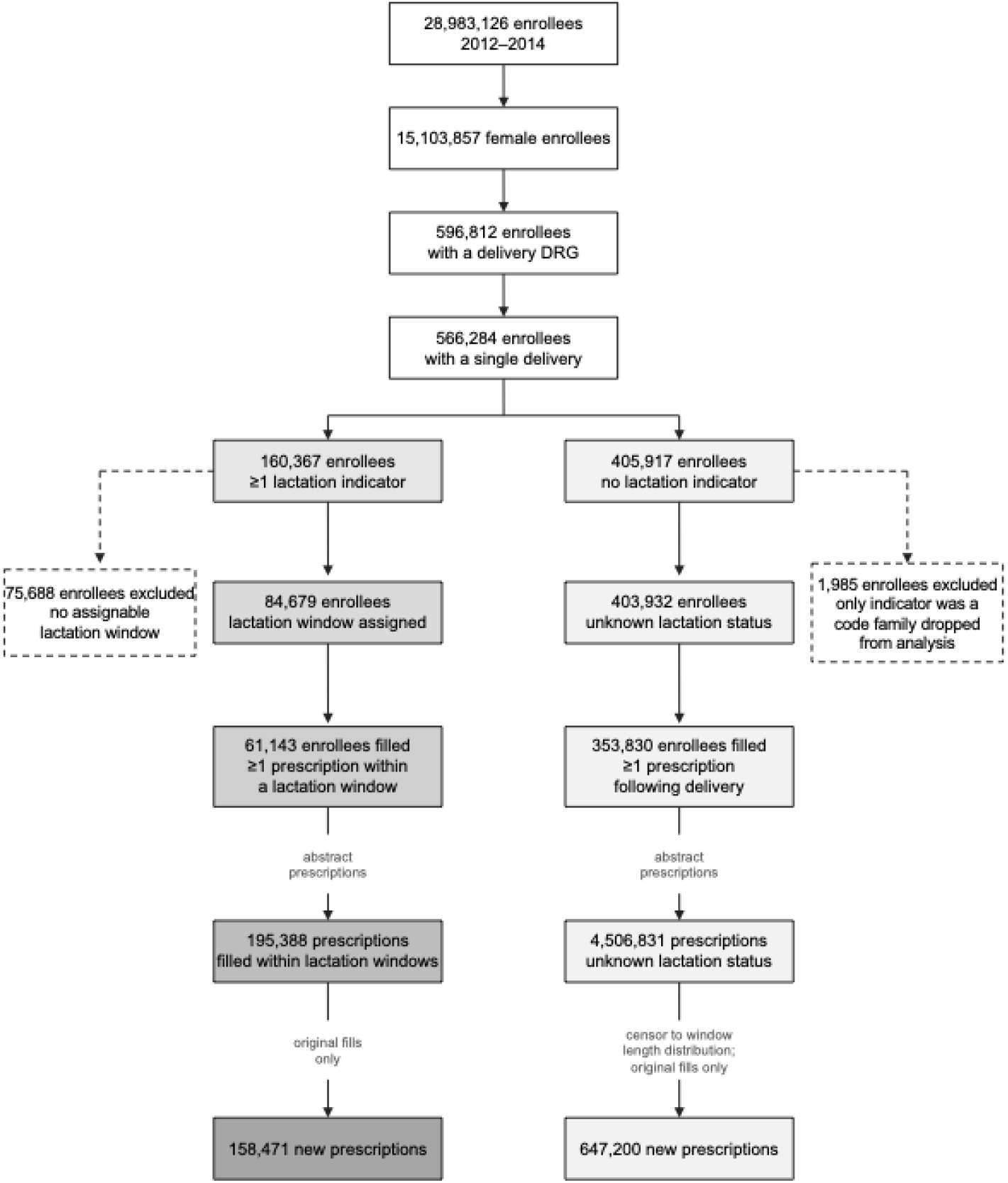
Identification of Postpartum Enrollees, Lactation Windows, and Prescriptions in the Merative MarketScan Commercial Claims Database, 2012-2014. Postpartum enrollees were identified by delivery-related MS-DRG and restricted to those with a single delivery during the study period. A lactation window ran from the delivery date to the date of the last surrogate lactation indicator. Enrollees with at least one lactation indicator did not all yield an assignable window such as acquisition of a breast pump during the prenatal period, and these enrollees were excluded. Women whose only lactation indicator belonged to a code family dropped during analysis were also excluded. New prescriptions are original fills, identified by the refill number recorded in the outpatient prescription file. Windows for the unknown lactation status group were assigned by superimposing the delivery-year-specific distribution of observed lactation window lengths. MS-DRG indicates Medicare Severity Diagnosis Related Group.

**Figure 2.**
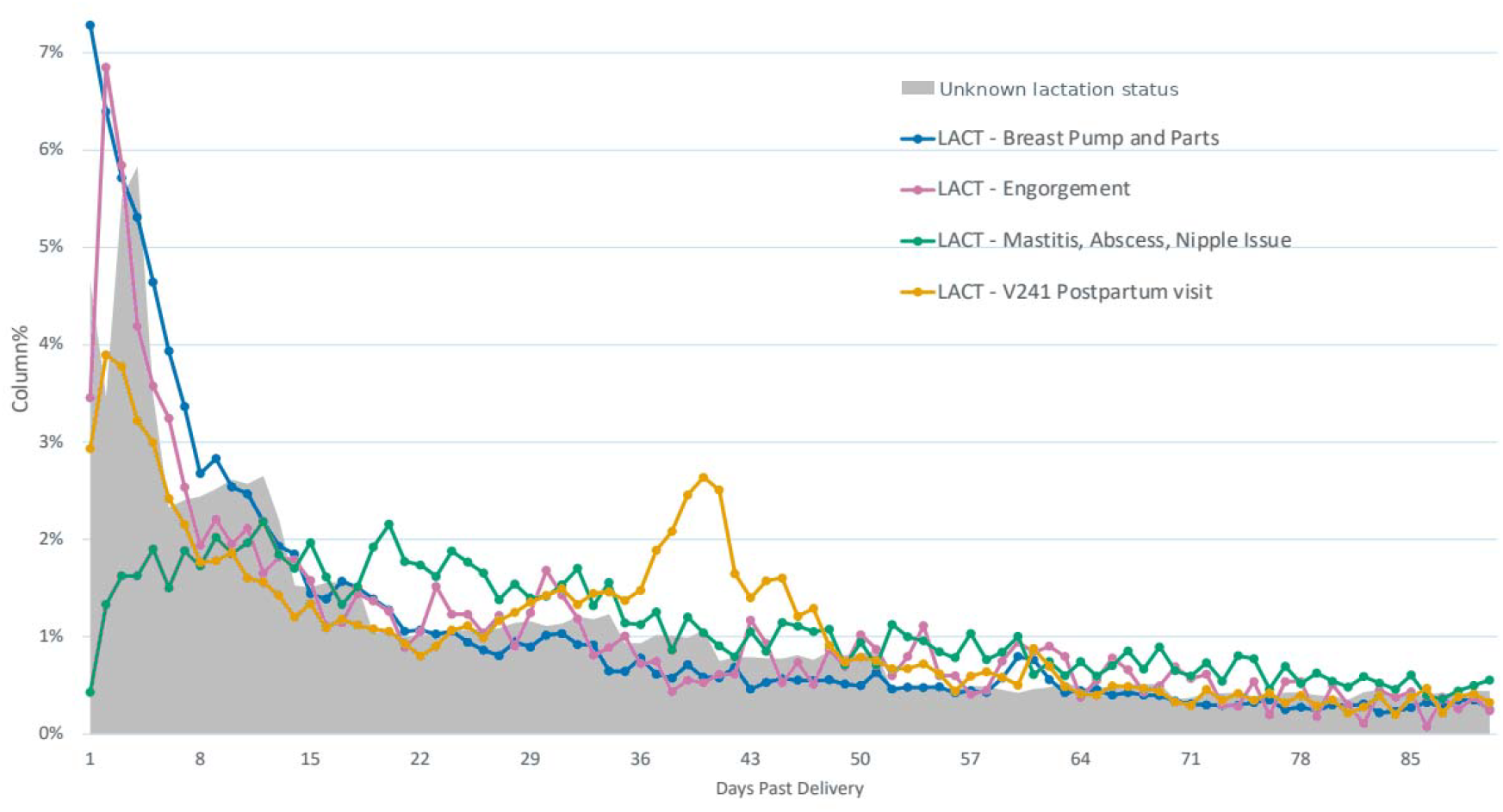
Timing of Prescription Fills During the First 90 Days After Delivery, by Lactation Status and Last Lactation Indicator. Values are the percentage of each group’s prescriptions filled on a given day, so each series sums to 100% across the period shown; the figure describes the within-group distribution of fill timing rather than the probability of a fill. Shaded area, postpartum enrollees of unknown lactation status; lines, prescriptions filled within lactation windows, stratified by the indicator that closed the window. Claims for breast pumps and supplies cluster in the first two weeks, and fills associated with the postpartum visit code peak near six weeks, consistent with routine obstetric follow-up. Series colors were selected for legibility under common forms of color vision deficiency.

Differences in patient characteristics by lactation status were statistically significant across all variables examined (Table 2). Women with an identified lactation window were older at the time of dispensing, with a mean (SD) age of 31.2 (5.02) years compared with 30.5 (5.68) years, and filled new prescriptions later, at a median (IQR) of 2 (0-34) days after delivery compared with 1 (0-22) day. Prescriptions originated disproportionately from the Southern US in both groups, more so among women of unknown lactation status (49.2%) than among lactating women (39.4%). Median day supply was longer during lactation, at 10 (IQR 5-20) days versus 8 (IQR 5-25) days, and medications for acute conditions accounted for 51.9% of prescriptions during lactation and 49.5% in the unknown group.

**Table 2.** Characteristics of postpartum enrollees by lactation status.

| Parameter | Lactation window<br>(N=84,679) | Unknown status<br>(N=403,932) | P |
| --- | --- | --- | --- |
| Age at delivery, mean (SD) | 29.59 (5.08) | 30.56 (5.86) | <0.0001 |
| Mode of delivery |  |  | <0.0001 |
| Cesarean w CC/MCC (765) | 13,578 (16.0%) | 58,635 (14.5%) |  |
| Cesarean w/o CC/MCC (766) | 15,487 (18.3%) | 86,171 (21.3%) |  |
| Vaginal w sterilization/D&C (767) | 578 (0.7%) | 5,020 (1.2%) |  |
| Vaginal w OR procedure (768) | 134 (0.2%) | 582 (0.1%) |  |
| Vaginal w complicating dx (774) | 8,267 (9.8%) | 35,225 (8.7%) |  |
| Vaginal w/o complicating dx (775) | 46,627 (55.1%) | 218,299 (54.1%) |  |
| Number of Prescriptions |  |  | <0.0001 |
| Prescriptions: 0 | 26,620 (31.4%) | 148,814 (36.9%) |  |
| Prescriptions: 1–4 | 50,807 (60.0%) | 230,050 (57.0%) |  |
| Prescriptions: 5–10 | 6,362 (7.5%) | 21,686 (5.4%) |  |
| Prescriptions: >10 | 891 (1.1%) | 3,382 (0.8%) |  |
| Footnote: Percentages are of the total; counts for nine enrollees with other delivery DRGs are not shown. P values are from the chi-square test for categorical variables and the Mann-Whitney U test for age, which was not normally distributed. Prescription counts are of all prescriptions filled after delivery within the assigned window. CC indicates complication or comorbidity; D&C, dilation and curettage; MCC, major complication or comorbidity; MS-DRG, Medicare Severity Diagnosis Related Group; SD, standard deviation. |  |  |  |

Prescription preferences were evident at the level of therapeutic group and class. Central nervous system agents were the most prevalent group in both, though less so during lactation (54.5% versus 63.1%), while anti-infectives were more prevalent during lactation (17.5% versus 11.0%). Gastrointestinal drugs (2.7% versus 2.1%) and vitamin products (2.4% versus 1.9%) were also more common during lactation, and cardiovascular agents slightly less common (3.0% versus 3.4%). Within classes the pattern was more specific. Combination analgesic and opioid agonist products were dispensed at identical rates in the two groups (50.0% each), but nonsteroidal anti-inflammatory drugs were more common during lactation (41.8% versus 39.8%), while antidepressants, benzodiazepines, anxiolytics and sedatives, amphetamine stimulants, anticonvulsants, and antipsychotics were all less common. The largest divergence was hormonal: progestins (59.5% versus 41.6%) and thyroid hormones (16.2% versus 11.1%) predominated during lactation, whereas combined oral contraceptives accounted for 8.2% of hormonal prescriptions during lactation against 29.0% in the unknown group.

The thirty most frequently dispensed new prescriptions during lactation are presented in Table 3. Ibuprofen accounted for 21.5% of all new fills, followed by combinations of acetaminophen with oxycodone, hydrocodone, or codeine, consistent with the stepwise approach recommended for postpartum pain.^18^ Norethindrone, a progestin-only contraceptive preferred during lactation because it is not thought to interfere with milk production, ranked fourth.^19^ Cephalexin, dicloxacillin, sulfamethoxazole-trimethoprim, and clindamycin are consistent with treatment of lactational mastitis,^20^ and sertraline with postpartum depression and anxiety.

**Table 3.**
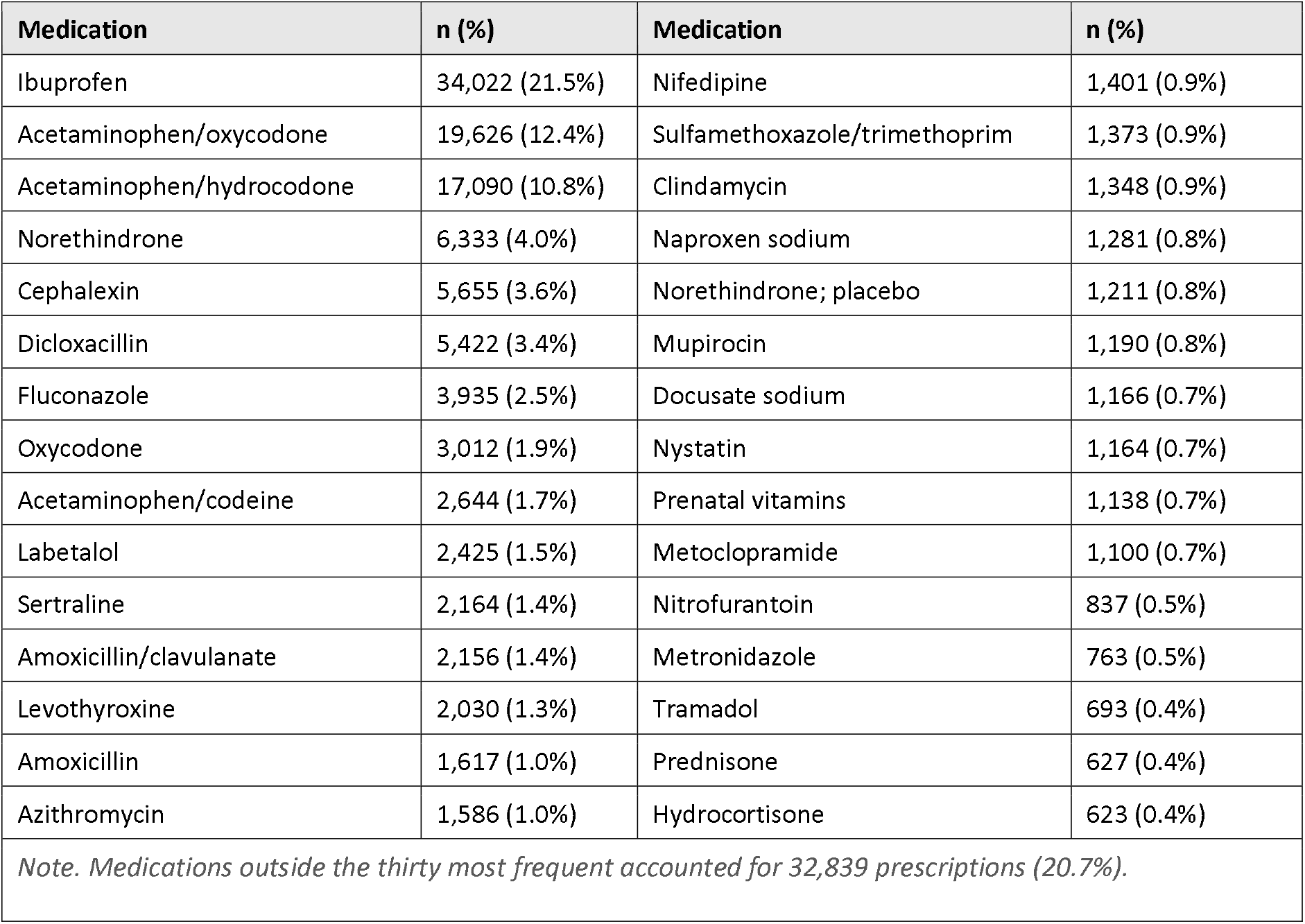
Thirty most frequently dispensed new prescriptions filled within lactation windows (N = 158,471)

| Medication | n (%) | Medication | n (%) |
| --- | --- | --- | --- |
| Ibuprofen | 34,022 (21.5%) | Nifedipine | 1,401 (0.9%) |
| Acetaminophen/oxycodone | 19,626 (12.4%) | Sulfamethoxazole/trimethoprim | 1,373 (0.9%) |
| Acetaminophen/hydrocodone | 17,090 (10.8%) | Clindamycin | 1,348 (0.9%) |
| Norethindrone | 6,333 (4.0%) | Naproxen sodium | 1,281 (0.8%) |
| Cephalexin | 5,655 (3.6%) | Norethindrone; placebo | 1,211 (0.8%) |
| Dicloxacillin | 5,422 (3.4%) | Mupirocin | 1,190 (0.8%) |
| Fluconazole | 3,935 (2.5%) | Docusate sodium | 1,166 (0.7%) |
| Oxycodone | 3,012 (1.9%) | Nystatin | 1,164 (0.7%) |
| Acetaminophen/codeine | 2,644 (1.7%) | Prenatal vitamins | 1,138 (0.7%) |
| Labetalol | 2,425 (1.5%) | Metoclopramide | 1,100 (0.7%) |
| Sertraline | 2,164 (1.4%) | Nitrofurantoin | 837 (0.5%) |
| Amoxicillin/clavulanate | 2,156 (1.4%) | Metronidazole | 763 (0.5%) |
| Levothyroxine | 2,030 (1.3%) | Tramadol | 693 (0.4%) |
| Amoxicillin | 1,617 (1.0%) | Prednisone | 627 (0.4%) |
| Azithromycin | 1,586 (1.0%) | Hydrocortisone | 623 (0.4%) |
| *Note. Medications outside the thirty most frequent accounted for 32,839 prescriptions (20.7%).* |  |  |  |

Among prescriptions filled during lactation windows, 35.4% were for medications classified at the time as L1, 15.8% L2, and 43.7% L3; 0.34% were L4 and 0.14% L5. Published lactation data were characterized as limited for 52.0% of prescriptions and absent for 42.4%. The corresponding proportions in the unknown lactation status group were 34.5% and 38.7%, with no lactation risk category available for 25.0% of prescriptions compared with 4.7% during lactation.

Provider type was linked for 32,616 prescriptions during lactation (20.6%) and 98,985 in the unknown group (15.3%). Obstetricians were the most frequent prescribers in both groups but accounted for only about one third of prescriptions during lactation and one quarter in the unknown group. Family practice providers were second, responsible for approximately 20% of postpartum prescriptions, followed by internal medicine and emergency medicine. Because licensure and specialty were recorded in a single variable, the two could not be separated; physicians were linked most often, followed by nurse practitioners and physician assistants, and then midwives, with midwife-linked prescriptions more common during lactation. Place of service was linked for 32,499 and 98,633 prescriptions respectively and was overwhelmingly an office or clinic visit (82.5% during lactation, 79.5% unknown).

## 4. Discussion

Among 566,284 commercially insured women with a single delivery, 84,679 (15%) could be assigned a lactation window from indicators already present in claims, and 158,471 new prescriptions were filled within those windows. Windows were short (median 17 days) and closed most often on a breast pump claim, so the method observes the early postpartum weeks most completely. What was dispensed was the medication of routine postpartum care: ibuprofen alone accounted for a fifth of new fills, and central nervous system agents, anti-infectives, and hormonal products together made up more than 80% of the total. Relative to women of unknown lactation status, dispensing during lactation favored progestins over combined oral contraceptives as well as drugs with at least some published lactation data, yet 42% of fills were for medications with no lactation data at the time. Obstetricians wrote only about a third of these prescriptions.

Lactation status is not recorded in administrative claims. Investigators have relied on data internal to a single health system,^10^ purpose-built research cohorts,^11^ prospective registries,^12^ or systems that capture infant feeding directly, whether a health record flowsheet^8^ or a national breastfeeding register.^9^ Each depends on someone having recorded lactation. The method described in this current study does not: it infers lactation from indicators already present in claims. When evaluating medications prescribed during lactation in the current study, they overlap substantially with those reported by other studies, with contraception, analgesics, anti-infectives, thyroid hormones, and antidepressants recurring across methods and countries.^8-9, 21^

The prescribers were less predictable. Obstetric and perinatal specialists are typically assumed to manage medication during lactation, but they wrote only about a third of prescriptions filled during lactation windows, and this was the early postpartum period when their involvement should be greatest. Family practice wrote roughly a fifth; internal medicine and emergency medicine both appear; psychiatric prescribers barely register despite maternal mental health being a stated national priority.^22^ Prescribing itself was conservative, favoring medications with published lactation data and rarely reaching drugs categorized as potentially hazardous to the breastfed infant, consistent with the risk avoidance described in earlier qualitative work^23^ and with the low rate of discordant prescribing reported in Brazil.^21^

Selecting drugs for lactation pharmacokinetic research has necessarily depended on expert judgment, because national utilization data have not existed. The prioritization process itself acknowledged this, calling for claims- and EHR-based description of medication use during lactation as the step preceding drug-specific safety and pharmacokinetic study.^3^ Just over half of prescriptions filled during lactation windows were for medications with some published lactation data and 42% for medications with none; among postpartum women of unknown lactation status the proportions reversed. These data cannot establish the direction of that association, and drug age and general-population use plausibly contribute to both dispensing frequency and evidence availability. A previous study of Canadian providers surveyed showed that half of them reported at least one medication class they were not confident counseling on, and only a quarter always asked whether a patient was breastfeeding when discussing a medication.^24^

Some studies also show that patients also experience some apprehension in taking medication when they are lactating. Asked to rate their comfort using a prescription medication while nursing, 79.1% of Canadian parents were uncomfortable or very uncomfortable when no studies of infant exposure exist on the prescribed medication, and more than half reported deliberately not starting a medication while breastfeeding.^24^ Comparable proportions appear elsewhere: 62.2% of Norwegian women with migraine abstained from medication because of breastfeeding on at least one occasion,^25^ and intentional underdosing or non-use was the leading cause of suboptimal treatment effect in a consultation cohort of lactating women.^6^ Because claims record dispensed fills rather than prescribing decisions, medications declined before reaching a pharmacy or left at the pharmacy by a patient are absent from these data, and they are unlikely to be absent at random. In support of that, recent evidence shows that when asked what would make them comfortable, 67.6% of parents named a medication that had been taken by many people without evidence of adverse effects in nursing infants, and providers answered the same way, with 58.6% comfortably recommending a medication on those grounds.^24^ MarketScan links infants to the primary beneficiary, so a woman identified as lactating can be linked to her infant, and the medications identified here can be carried into pharmacoepidemiologic analysis of adverse events among breastfed infants.

The algorithm developed to define lactation is specified for commercial claims. Medicaid, which finances approximately four in ten U.S. births,^26^ would require substantial adaptation rather than direct application: because benefits are set at the state level, the surrogate indicators do not carry equivalent meaning across states. Breast pump coverage in particular varies in scope, in prior authorization and medical necessity requirements, and in whether pumps are billed as durable medical equipment or through the pharmacy benefit, while pumps supplied through WIC generate no claim at all,^27^ a consequential gap given that pump claims closed over half of the windows identified here. Postpartum eligibility was limited to 60 days in most states before 2022,^28^ censoring windows administratively rather than biologically, and delivery identification by MS-DRG may not transfer where states pay on other bases. Investigators applying this method to Medicaid data should characterize indicator composition by state before interpreting any substantive result. Other extensions are more readily available. The phenotype can be applied to ICD-10-era commercial data using code equivalents. More substantially, because MarketScan links dependents to the primary beneficiary, a woman identified as lactating can be linked to her infant, making it possible to study infant outcomes following maternal medication exposure through human milk in a national population. Recording lactation directly has been a stated federal priority since PRGLAC recommendation 12,^29^ and lactation data elements have since been added to the USCDI+ Maternal Health dataset,^30^ but they are exploratory rather than required for certified records, are collected prospectively, and do not appear in claims databases. Until lactation is both mandated and linkable to dispensing data, inference from surrogate indicators remains necessary.

The major strength of the current study is the use of a large national database with well characterized information of medication use during the postpartum period. The method developed to identify lactation windows is fairly simple and straightforward; it requires no natural language processing, no access to clinical free text, and no machine learning infrastructure. The following limitations should be considered. The phenotype has not been validated against a chart-confirmed reference standard. Unquantified misclassification of lactation status is characteristic of current work in this area. Selection bias is likely, as women who lactated but do not have the surrogate indicators to identify lactation may display different prescription characteristics. Claims for breast pumps and supplies closed 53.2% of lactation windows, so mothers who express milk are over-represented, and pump acquisition is itself patterned by insurance coverage and by return-to-work expectations. Lactation windows measure a minimum duration of lactation rather than its true duration, since a window closes at the last observed indicator rather than at weaning. Windows were also concentrated early. This limits what can be said about medication use later in the first postpartum year. It also means the method observes most completely the interval in which pharmacologic evidence is thinnest and infants are at highest exposure risks, between delivery and six weeks postpartum.^1^

The study lacks a non-lactating comparison group. The unknown lactation status group contains both lactating and non-lactating women, which makes it difficult to establish whether the dispensing patterns observed are specific to lactation. Claims capture dispensed fills rather than prescribing decisions, and a filled prescription does not establish that the medication was taken.

These data describe medications dispensed between 2012 and 2014 and should be read as a historical account. The study period predates the ICD-10 transition and prescribing practices have changed since. This limitation applies to the descriptive findings rather than to the algorithm used to define lactation Finally, findings of the current study are generalizable only to commercially insured individuals.

## 5. Conclusion

Lactation can be identified in administrative claims from indicators already present in the data, which makes it possible to describe what lactating women are dispensed at national scale. What emerges is clinically coherent, conservative, and spread across many prescriber specialties rather than concentrated in obstetrics. Two steps follow directly. The phenotype should be applied to and validated for contemporary ICD-10 data, for which the code equivalents are provided, so that current practice can be described. Those results would inform which commonly dispensed medications warrant pharmacoepidemiologic analysis of adverse events in the breastfed infant. Infant safety for breastfeeding medication exposures is the question mothers are asking, and it is now within grasp.

## Data Availability

Data availability. The MarketScan data are licensed and cannot be redistributed. Complete code lists and the phenotype specification are provided as Supplementary Material. Reasonable requests for analytic code will be considered by the corresponding author.
Data license and ownership. Certain data used in this study were supplied by International Business Machines Corporation as part of one or more Merative MarketScan Research Databases. Any analysis, interpretation, or conclusion based on these data is solely that of the authors and not International Business Machines Corporation.

## Acknowledgments

**Acknowledgments**

The authors thank Joseph Bocchino, EdD, and Anthony Scialli, MD, for their guidance on the doctoral work from which this analysis derives.

## AI Statement

The authors acknowledge the use of Claude (Anthropic, 2026) to assist with manuscript preparation. The authors reviewed and verified all AI-assisted output, including numerical values and references, and take full responsibility for the content of this article.

## Author Contributions

CRediT: Conceptualization: KK; Data curation: KK; Formal Analysis: KK, DA; Funding acquisition; Methodology: KK, DA; Supervision: DA; Visualization: KK; Writing – original draft: KK; Writing – review & editing: KK, DA

## Statements and Declarations

### Ethical Considerations

Deidentified data; not human subjects research. A data management plan was approved by The George Washington University data librarian prior to analysis.

### Consent to Participate

Not applicable Consent for Publication. Not applicable

### Prior publication

This work derives from the corresponding author’s doctoral dissertation, The George Washington University, 2024.

## Data Statement

### Data availability

The MarketScan data are licensed and cannot be redistributed. Complete code lists and the phenotype specification are provided as Supplementary Material. Reasonable requests for analytic code will be considered by the corresponding author.

### Data license and ownership

Certain data used in this study were supplied by International Business Machines Corporation as part of one or more Merative MarketScan Research Databases. Any analysis, interpretation, or conclusion based on these data is solely that of the authors and not International Business Machines Corporation.

