## Supplemental Materials for "No Field for Lactation: Inferring Lactation Status From US Commercial Claims to Describe Postpartum Medication Dispensing"

### Supplementary Material

#### Table S1. Delivery identification — MS-DRG codes

| **MS-DRG (FY2014)** | **Description** |
| --- | --- |
| 765 | Cesarean section with CC/MCC |
| 766 | Cesarean section without CC/MCC |
| 767 | Vaginal delivery with sterilization and/or D&C |
| 768 | Vaginal delivery with O.R. procedure except sterilization and/or D&C |
| 774 | Vaginal delivery with complicating diagnoses |
| 775 | Vaginal delivery without complicating diagnoses |

*Note. Obstetric MS-DRGs were restructured effective FY2018. Investigators applying this approach to data from October 2017 onward should identify the corresponding codes in the CMS MS-DRG Definitions Manual for the fiscal year of their data.*

#### Table S2. Lactation phenotype — surrogate indicators

| **Indicator** | **ICD-10-CM / ICD-9-CM / CPT / HCPCS** |
| --- | --- |
| Nipple infection | ICD-10 O91.03 (AWL); ICD-9 675.04 (AWPP) |
| Breast abscess | ICD-10 O91.13 (AWL); ICD-9 675.14 (AWPP) |
| Non-purulent mastitis | ICD-10 O91.23 (AWL); ICD-9 675.82, 675.84 (AWPP) |
| Retracted nipple | ICD-10 O92.03 (AWL); ICD-9 676.02, 676.04 (AWPP) |
| Cracked nipple | ICD-10 O92.13 (AWL); ICD-9 676.12, 676.14, 676.34 (AWPP) |
| Galactorrhea | ICD-10 O92.6; ICD-9 676.62, 676.64 |
| Breast engorgement ᵃ | ICD-10 O92.29 (other disorders); ICD-9 676.24 ᵇ |
| Delayed lactation ᵃ | ICD-9 676.84 (other disorders) ᵇ |
| Supervision of lactating mother | ICD-10 Z39.1; ICD-9 V24.1 |
| Candidiasis, breast or nipple ᵃ | ICD-10 B37.89 (location: other); ICD-9 112.89 (location: other) ᵇ |
| Hypogalactia ᵃ | ICD-10 O92.4; no ICD-9 equivalent |
| Lactation class | CPT S9443 |
| Breast pump or supplies | HCPCS E0602, E0603, E0604, A4281, A4282, A4283, A4284, A4285, A4286, K1005 |

*AWL, associated with lactation; AWPP, associated with postpartum condition or complication.*

*ᵃ May not be helpful in identifying clinically relevant lactating behavior. ᵇ Not used in final analysis.*
